# HYPE: Predicting Blood Pressure from Photoplethysmograms in a Hypertensive Population

**DOI:** 10.1101/2020.05.27.20107243

**Authors:** Ariane M. Sasso, Suparno Datta, Michael Jeitler, Nico Steckhan, Christian S. Kessler, Andreas Michalsen, Bert Arnrich, Erwin Böttinger

**Author notes:** The two authors contributed equally to this paper (Ariane M. Sasso, Suparno Datta).

## Abstract

The state of the art for monitoring hypertension relies on measuring blood pressure (BP) using uncomfortable cuff-based devices. Hence, for increased adherence in monitoring, a better way of measuring BP is needed. That could be achieved through comfortable wearables that contain photoplethysmography (PPG) sensors. There have been several studies showing the possibility of statistically estimating systolic and diastolic BP (SBP/DBP) from PPG signals. However, they are either based on measurements of healthy subjects or on patients on intensive care units (ICUs). Thus, there is a lack of studies with patients out of the normal range of BP and with daily life monitoring out of the ICUs. To address this, we created a dataset (HYPE) composed of data from hypertensive subjects that executed a stress test and had 24-hours monitoring. We then trained and compared machine learning (ML) models to predict BP. We evaluated handcrafted feature extraction approaches vs image representation ones and compared different ML algorithms for both. Moreover, in order to evaluate the models in a different scenario, we used an openly available set from a stress test with healthy subjects (EVAL). The best results for our HYPE dataset were in the stress test and had a mean absolute error (MAE) in mmHg of 8.79 (±3.17) for SBP and 6.37 (±2.62) for DBP; for our EVAL dataset it was 14.74 (±4.06) and 7.12 (±2.32) respectively. Although having tested a range of signal processing and ML techniques, we were not able to reproduce the small error ranges claimed in the literature. The mixed results suggest a need for more comparative studies with subjects out of the intensive care and across all ranges of blood pressure. Until then, the clinical relevance of PPG-based predictions in daily life should remain an open question.

## 1 Introduction

According to the Global Disease Burden (GBD) study, high blood pressure (BP) (i. e. hypertension) is the risk factor that leads to more deaths worldwide [16]. The standard way of monitoring this condition is through the measurement of BP using an uncomfortable cuff-based device [24]. Fortunately, comfortable and common wearables can already detect changes in the flow of blood through a photoplethysmography (PPG) sensor [1]. The PPG signal (photoplethysmogram) obtained from it is already used with success to estimate heart rate [19] and, has the potential to go beyond that into accurate BP prediction [5,2].

Most of the work in this area focus on building predictive models for patients in intensive care units (ICUs) [12,23,20]. However, data collected from regular life contain motion artifacts that are not observed in intensive care. Additionally, models that work on healthy populations [17,18] should also be validated on hypertensive populations for guarantying their applicability in BP monitoring. Hence, in our work we focused on assembling a dataset containing data from subjects with hypertension (HYPE) during a stress test and 24-hours monitoring.

We then evaluated machine learning (ML) models for predicting BP from PPG in the HYPE dataset and also in a dataset from healthy subjects during a stress test (EVAL). From the PPG signals, we extracted features from the time domain plus their image representations. Errors as low as the ones in the literature—for patients in the ICU or healthy subjects—could not be reproduced, even after processing the PPG signals with diverse time windows and filters.

This work is detailed as follows: section 2 shows previous work in the field and section 3 describes the datasets and methods we used to predict BP from the PPG signal. In section 4 we convey our findings and results, followed by a discussion in section 5 and section 6 describing the implications of this work.

## 2 Related Work

Existing work focuses on predictive models using MIMIC [7], a dataset that contains physiological signals including PPG and ambulatory BP (ABP) from patients in ICUs. Kurylayak et al. [12] and Wong et al. [23] have both applied artificial neural networks (ANN) to predicted BP in this dataset and reported success. However, they used unknown or small sample sizes as can be seen in Table 1. Moreover, Kurylayak et al. only extracted time domain features from the PPG signal while Wong et al. also extracted frequency domain ones. Conversely, Slapničar et al. [20] tried a spectro-temporal ResNet with all features in a larger sample size but could not report the same success as his predecessors.

**Table 1:** Blood Pressure Prediction from Photoplethysmograms

| WORK | DATASET | FEATURES<br>(PPG) | METHOD | MAE<br>(mmHg) |
| --- | --- | --- | --- | --- |
| [12] | MIMIC<br>15000<br>pulsations | Time<br>Domain | ANN | SBP 3.80 ($\pm 3.46$)<br>DBP 2.21 ($\pm 2.09$) |
| [23] | MIMIC<br>72 subjects | Time and<br>Frequency<br>Domain | ANN | SBP 4.02 ($\pm 2.79$)<br>DBP 2.27 ($\pm 1.82$) |
| [20] | MIMIC<br>510 subjects | Time and<br>Frequency<br>Domain | Spectro-Temporal<br>ResNet | SBP 9.43 (N/A)<br>DBP 6.88 (N/A) |
| [17] | Healthy Subjects<br>Daily Life<br>22 subjects | Time and<br>Frequency<br>Domain | Ensemble of<br>Regression<br>Trees | SBP 6.70 (N/A)<br>DBP 4.42 (N/A) |
| [18] | Healthy Subjects<br>Controlled<br>50 subjects | Time<br>Domain | ANN | SBP 4.1 (N/A)<br>DBP 1.7 (N/A) |

Others have tried to collect data from healthy subjects in daily life such as Lustrek et al. [17]. They have used the device empatica E4^†^ and evaluated a range of machine learning (ML) techniques, achieving the best results with an ensemble of regression trees and the leave-one-subject-out (LOSO) validation strategy. However, they had to use ground truth BP from each subject to personalize the algorithm. Lastly, there is the work of Manamperi et al. [18], in which they evaluated ANN in MIMIC and in a set with data from voluntary subjects (assumed as healthy). They claim to have done the second evaluation in a non-clinical scenario, but the subjects were mainly at rest in their experiment.

Therefore, the current state-of-the-art does not give yet conclusions about the use of PPG to predict BP in diverse populations and in daily life. There is a clear need for more comparative studies both with healthy and hypertensive subjects and in different scenarios, especially outside of controlled conditions.

## 3 Methods

### 3.1 Datasets

In our work we used two datasets: one created by us with data from a hypertensive population (HYPE) and one that is openly available containing data from a healthy population (EVAL). It should be noted that both datasets recorded patients during a stress test and HYPE also during 24-hours monitoring. We describe the two datasets below.

> **HYPE** This dataset was created by us as part of the CardioVeg study (NCT03901183) approved by the Ethics Committee from Charité, Berlin (no. EA4/025/19). Data was collected from 12 subjects (6 female) in the age range of 31-75 (median 60) that had hypertension. The study collected data from a stress test (1) and from 24-hours monitoring (2) using the empatica E4 wristband as the PPG source and the Spacelabs (SL 90217) BP monitor.
>
> (1) **Stress Test**. The subjects followed a protocol in which they watched a relaxing video for 5 minutes then had their BP taken by a physician five times with an interval of 1 minute per measurement [24]. Then, the patients biked in an ergonomic bike from 5 to 10 minutes and relaxed again. During the second relaxation phase their BP was measured again 5 times with a 1 minute interval. This dataset contains a total of 95 BP recordings. One subject could not bike due to extreme high BP and another one had a failure in the wearable device. Therefore, this experiment had 10 subjects (5 female).
>
> (2) **24 Hours**. In this phase, the same subjects from the stress test were monitored for 24-hours during regular day activities. The Spacelabs monitoring device was configured to measure BP every 30 minutes during the day and every hour during the night. This dataset contains a total of 464 BP recordings and all 12 subjects were measured.
>
> **EVAL** This dataset was generated by Esmaili et al. [3]. The original paper tried to estimate BP based on pulse transit time (PTT) and pulse arrival time (PAT). Both variables are derived from the differences between the PPG and ECG signals. This data was collected from 26 healthy subjects in the age range of 21–50 years. The subjects were required to run for 3 minutes at the speed of 8 km/h to induce perturbations in their BP values. Directly after the exercise the subjects were made to sit upright and BP values were measured along with PPG and ECG. A force-sensing resistor (FSR) was used under the BP monitor cuff to measure the instantaneous cuff pressure. With the FSR it was possible to pin point the exact time when the SBP and DBP were measured. A total of 152 BP values were recorded in this dataset.

### 3.2 Handcrafted Feature Extraction Methods

Our first approach entailed extracting handcrafted features from the PPG signal. Time windows of 15, 30 and 45 seconds around the BP measurement were used for our experiments. To eliminate motion artefacts induced by wrist movements sections in which the Euclidean norm of x-, y- and z-acceleration lied outside of an interval of 25% of the standard deviation around the sample mean, for the current window, were removed from consideration. The motion removal was only done for the HYPE dataset as the EVAL dataset did not contain any motion signals corresponding to the PPG recording. We also experimented with signal normalization and filters such as Chebyshev II and Butterworth, since they were reported as the best filters for PPG signals [15]. For the processed signal, the PPG cycles were then identified with a standard peak detection function.

All detected cycles in the same window were combined into a custom PPG signal template (details in section B), following a procedure described by Li and Clifford [13]. Individual cycles were then compared with the template using two signal quality indices (SQI): (1) direct linear correlation and (2) direct linear correlation between the cycle, re-sampled to match the template length, and the template itself. Only if both correlations lied above 0.8, the cycle was further processed to extract features. This resulted in some BP intervals not having any features extracted since no cycles matching the template were identified.

After the clean PPG cycles have been identified, time domain features were extracted and the detailed list can be found in the Appendix (Table 4). The first step was to identify the first peak in the cycle, which corresponded to the systolic peak. Then for various percentages of the peak amplitude, we extracted the time between systolic peak and end of the cycle (*DW_n_*), start of the cycle and end of the cycle (*SW_n_* + *DW_n_*), and the ratio between the time in the cycle before and after the systolic peak (*DW_n_/SW_n_*). For every window, the mean and variance of each feature were computed and used as input for the models.

### 3.3 Image Representation Methods

An alternative approach to the manual feature extraction has recently gained much popularity involving convolutional neural networks (CNNs). The approach is to represent the waves as images and then use a transfer learning method based on pretrained CNNs to learn embedding from the images and use them to predict BP. The two different image-form representations of PPG signals that we tested were spectograms and scalograms, described below.

> **Scalograms**. A scalogram is usually plotted as a graph of time and frequency and it represents the absolute value of the Continuous Wavelet Transform (CWT) coefficients of a signal. The scalogram-CNN based approach was first discussed in Liang et al. [14]. However, it was only evaluated for hypertension stratification, not BP prediction. Before passing the signal to the CWT, we detrended it, i.e. subtracted the mean value from the input signal. CWT is a convolution of the input data sequence with a set of functions generated by the base wavelet. We used the complex Morlet wavelet function as the base wavelet, which is given by:
>
> 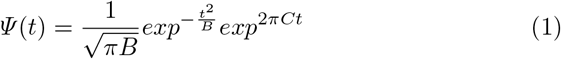
>
> The value of bandwidth frequency (B) and center frequency (C) was chosen to be 3 and 60 in the above equation, following the work of Liang et al. [14]. Compared to a spectrogram, a scalogram is usually better at identifying the low-frequency or fast-changing frequency component of the signal.
>
> **Spectograms**. A spectogram displays changes in the frequencies in a signal over time. A third dimension indicating the amplitude of a particular frequency at a particular time is represented by the intensity or color of each point in the plot. The spectogram-CNN approach to predict BP was first discussed in Slapnicar et al. [20]. Similar to scalograms, we detrended our signal before generating the spectogram plots. To generate a spectogram, digitally sampled signals in the time domain are broken up into windows, which usually overlap, and they are Fourier transformed to calculate the magnitude of the frequency spectrum for each window [21].

Figure 1 depicts a sample spectogram and scalogram generated from a PPG snippet of 15 seconds. The image representations of the signal were then fed into a ResNet architecture to learn the image embeddings [9]. The Residual Network or ResNet design enables us to train very deep neural networks without running into the vanishing gradient problem. Since our datasize is very small, instead of training a network from scratch, we decided to take a network which was already trained on the ImageNet dataset [8]. In particular, we used the ResNet18 architecture and took the embeddings from the penultimate layer of the network. We also experimented with Alexnet, but Resnet18 always performed marginally better [11]. This might be due to the fact the penultimate layer of the Resnet18 generates a 512 length embedding, whereas the AlexNet generates a embedding of length 4096. The larger size of the input vector, in spite of using feature selection and strong regularization techniques, might make it challenging for the models to learn from, due to the small data size.

**Fig. 1:**
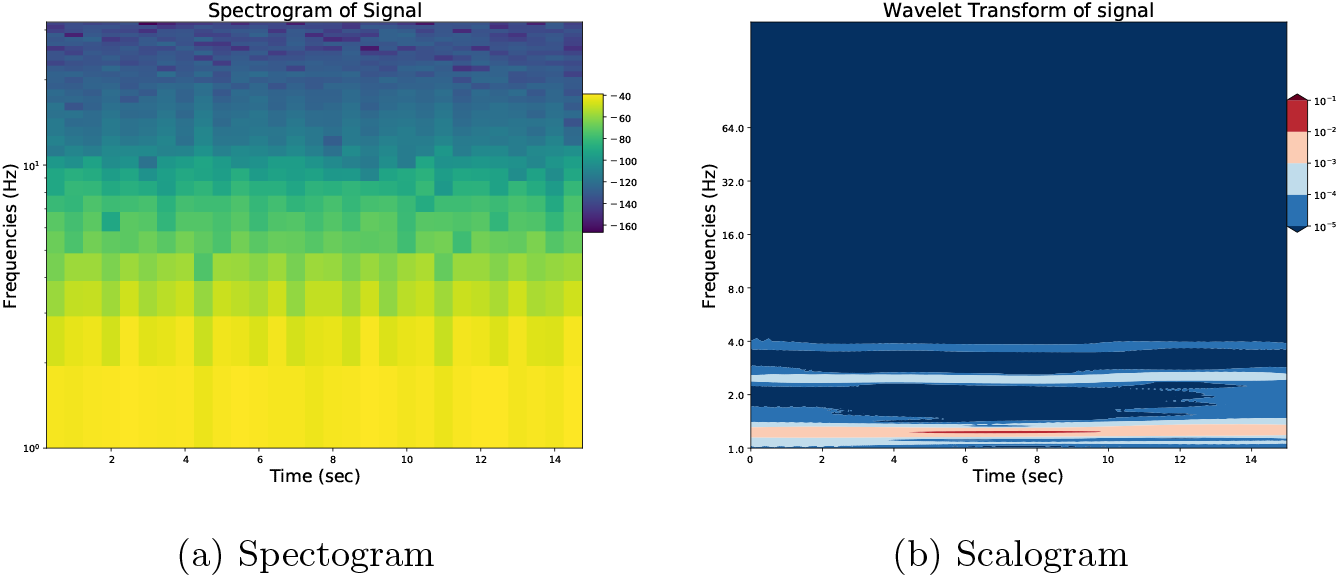
(a) displays a sample spectogram. (b) depicts a scalogram with the complex Morlet wavelet function used as the base wavelet for the same PPG signal.

### 3.4 Machine Learning Models

Previous works show that machine learning algorithms perform well in predicting BP from features derived from PPG and/or ECG. We have employed in our experiments three popular machine learning algorithms: (a) Generalised Linear Models (GLM) with Elastic Net regularisation [25], (b) Gradient Boosting Machines (GBM) [4], and (c) a recent more efficient implementation of GBMs called LightGBM (LGBM) [10] to predict the systolic and diastolic BP.

For prediction from the image embeddings, we used a Recursive Feature Elimination (RFE) technique with a support vector machine (SVM) with linear kernel as the base estimator, before pushing the vectors into the models.

### 3.5 Experimental Settings

In order to train models that are well generalizable, we used a leave-2-subjects-out cross validation for all our models, i.e. at every iteration we use data from 2 subjects as the test set, trained our models on the remaining data and repeated this procedure till all subjects have been at some point used as the test set. All hyper parameters were optimized empirically. We evaluated the models based on the mean absolute error (MAE). The MAE was calculated at each iteration and we calculated the mean and standard deviation of these values.

## 4 Results

In this section we report our experimental results. In Table 2 we show the comparison of the MAEs for predicting systolic blood pressure (SBP) between the different models in different datasets. Table 3 shows the results for diastolic blood pressure (DBP) prediction. The cells in these table contain the mean and standard deviation (in parenthesis) of the MAE of all cross validation folds. Noticeably, in the HYPE dataset feature extraction based methods consistently outperformed the image based methods. In EVAL the spectogram-representation based results outperformed the other two approaches. In both datasets, the best results for the spectogram based approach are usually marginally better than the best results of the scalogram based approach. For the image based methods the more advanced machine learning methods such as LGBM and GBM clearly outperformed the GLM model. This is most probably due to the comparatively large dimension of the input image embeddings. For the feature extraction based methods this difference is not so prominent, and in some cases the GLM turns out to be the best performing model. In general, based on the MAE values, predicting SBP appears to be more difficult than predicting DBP which is consistent with previous literature (see Table 1).

**Table 2:** MAE of the different models for SBP prediction in different datasets

| Dataset | Feature Selection |  |  | Spectograms-Resnet18 |  |  | Scalograms-Resnet18 |  |  |
| --- | --- | --- | --- | --- | --- | --- | --- | --- | --- |
|  | GLM | GBM | LGBM | GLM | GBM | LGBM | GLM | GBM | LGBM |
| EVAL | 16.71 | 16.19 | 16.00 | 17.59 | 15.24 | **14.74** | 16.71 | 15.68 | 15.46 |
| Stress Test | ($\pm 4.32$) | ($\pm 4.35$) | ($\pm 4.63$) | ($\pm 3.72$) | ($\pm 4.01$) | ($\pm 4.06$) | ($\pm 4.16$) | ($\pm 4.34$) | ($\pm 4.49$) |
| HYPE | 10.26 | **8.79** | 9.57 | 15.58 | 12.44 | 12.15 | 17.98 | 12.91 | 12.83 |
| Stress Test | ($\pm 1.18$) | ($\pm 3.17$) | ($\pm 1.65$) | ($\pm 1.21$) | ($\pm 2.72$) | ($\pm 2.72$) | ($\pm 2.11$) | ($\pm 2.57$) | ($\pm 2.63$) |
| HYPE | **14.44** | 14.83 | 14.74 | 18.71 | 16.92 | 17.07 | 18.97 | 17.03 | 17.30 |
| 24-hours | ($\pm 2.96$) | ($\pm 3.81$) | ($\pm 4.07$) | ($\pm 4.072$) | ($\pm 5.22$) | ($\pm 5.22$) | ($\pm 5.15$) | ($\pm 5.26$) | ($\pm 5.21$) |

**Table 3:** MAE of the different models for DBP prediction in different datasets

| Dataset | Feature Selection |  |  | Spectograms-Resnet18 |  |  | Scalograms-Resnet18 |  |  |
| --- | --- | --- | --- | --- | --- | --- | --- | --- | --- |
|  | GLM | GBM | LGBM | GLM | GBM | LGBM | GLM | GBM | LGBM |
| EVAL | 7.87 | 7.86 | 7.57 | 13.18 | **7.12** | 7.15 | 8.67 | 7.62 | 7.53 |
| Stress Test | ($\pm 2.07$) | ($\pm 2.27$) | ($\pm 2.38$) | ($\pm 13.01$) | ($\pm 2.32$) | ($\pm 2.42$) | ($\pm 3.08$) | ($\pm 2.35$) | ($\pm 2.48$) |
| HYPE | 7.50 | **6.37** | 7.22 | 11.98 | 9.55 | 9.52 | 12.18 | 9.51 | 9.35 |
| Stress Test | ($\pm 0.68$) | ($\pm 2.62$) | ($\pm 2.69$) | ($\pm 1.90$) | ($\pm 2.74$) | ($\pm 2.02$) | ($\pm 1.98$) | ($\pm 1.91$) | ($\pm 1.93$) |
| HYPE | 11.52 | **11.48** | 11.56 | 13.93 | 12.84 | 12.79 | 15.71 | 12.94 | 13.14 |
| 24-hours | ($\pm 3.05$) | ($\pm 3.57$) | ($\pm 2.03$) | ($\pm 3.20$) | ($\pm 4.00$) | ($\pm 4.10$) | ($\pm 3.83$) | ($\pm 4.00$) | ($\pm 4.03$) |

**Table 4:** Features extracted from PPG [12]. Here *n* ∊ {10, 25, 33, 50, 66, 75}

| Name | Description |
| --- | --- |
| SUT | Systolic Upstroke Time |
| DT | Diastolic Time |
| CP | Cardiac Period |
| $DW_n$ | Diastolic Width at $n\%$ amplitude |
| $SW_n + DW_n$ | Sum of Systolic Width and Diastolic Width at $n\%$ amplitude |
| $DW_n/SW_n$ | Ratio between Systolic Width and Diastolic Width at $n\%$ amplitude |

## 5 Discussion

### 5.1 Clinical Relevance

Cuff-less and continuous methods of measuring BP are particularly attractive as BP is one of the most important predictors of long term cardiovascular health [6]. Prediction models for BP based on PPG signals can be a very important stride in that direction. But for reliable continuous monitoring of BP, these models need to perform well during regular day-to-day activities and also for different patient populations. Apart from MIMIC there does exist a few other works that try to collect PPG and corresponding BP signals following a strict protocol (similar to HYPE: stress-test). To the best of our knowledge, our 24-hours dataset is the first attempt to collect this data from an uncontrolled environment where the subjects were free to do anything. Our evaluations do underline that it is indeed more challenging to accurately predict BP in such an uncontrolled environment.

### 5.2 Technical Relevance

In this work we impartially evaluated different models and approaches for BP prediction from PPG. Most of the methods have only been previously validated on MIMIC. Also, due to the large volume of existing work, very often the models were not compared against all available approaches. To the best of our knowledge, this is also the first work to compare the scalogram, spectogram and feature extraction approaches. We employ strong cross validation methods to make sure our results are robust. Our models and code are available openly to make sure this results can be reproduced and also applied to similar datasets when needed.

### 5.3 Limitations and Future Work

The major limitation of our work is related to the small size of the datasets we used. For that reason, it was not possible to train a deep Long Short Term Memory (LSTM) network, which in a few recent papers have demonstrated very promising results [22]. In future work, we would like to extend our dataset with more diverse patient populations and also with a longer observation period per patient. This will allow us to apply more data-demanding learning algorithms and, at the same time, to investigate how models trained in one population perform in a different one.

## 6 Conclusion

In conclusion, we presented a comprehensive comparison of different machine learning approaches to predict BP from PPG in two different datasets. We demonstrate that despite the plethora of work in this area, there exists a dearth of models that perform well in uncontrolled environments when the subjects indulge in various day-to-day activities. We show for small to medium sized datasets that feature extraction based methods can produce better results than the recent image based approaches. We hope our paper will induce more work that will look into the generalizability of these models.

## Data Availability

(EVAL): Openly Available Dataset
(HYPE): Waiting for Approval

https://www.kaggle.com/mkachuee/noninvasivebp

https://github.com/arianesasso/aime-2020

https://figshare.com/projects/AIME_2020/85166

## 7 Acknowledgements

We would like to thank Manisha Manaswini, Felix Musmann, Juan Carlos Nino Rodriguez, and Carolin Müller for their help during data collection and, also Harry Freitas da Cruz and Attila Wohlbrandt for giving many valuable insights.

## APPENDIX

## A Data and Code Availability

The code for the experiments is available at: https://github.com/arianesasso/aime-2020. Information on the HYPE dataset is also provided there. The EVAL dataset can be found at: https://www.kaggle.com/mkachuee/noninvasivebp.

## B Feature extraction

The features that were extracted from the PPG cycles are described in Table 4 and in Figure 2 following the work of Kurylyak et al. [12].

**Fig. 2:**
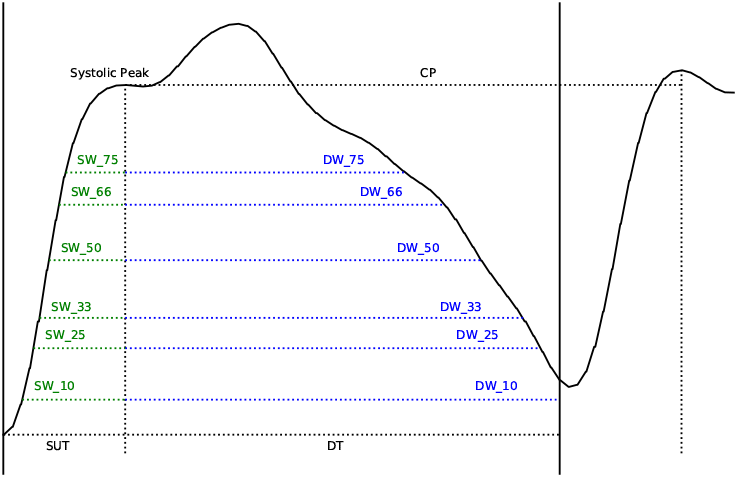
Extracted features from PPG

Figure 3a depicts the template used to detect valid cycles from the PPG signal and Figure 3b a set of valid cycles detected from the EVAL dataset.

**Fig. 3:**
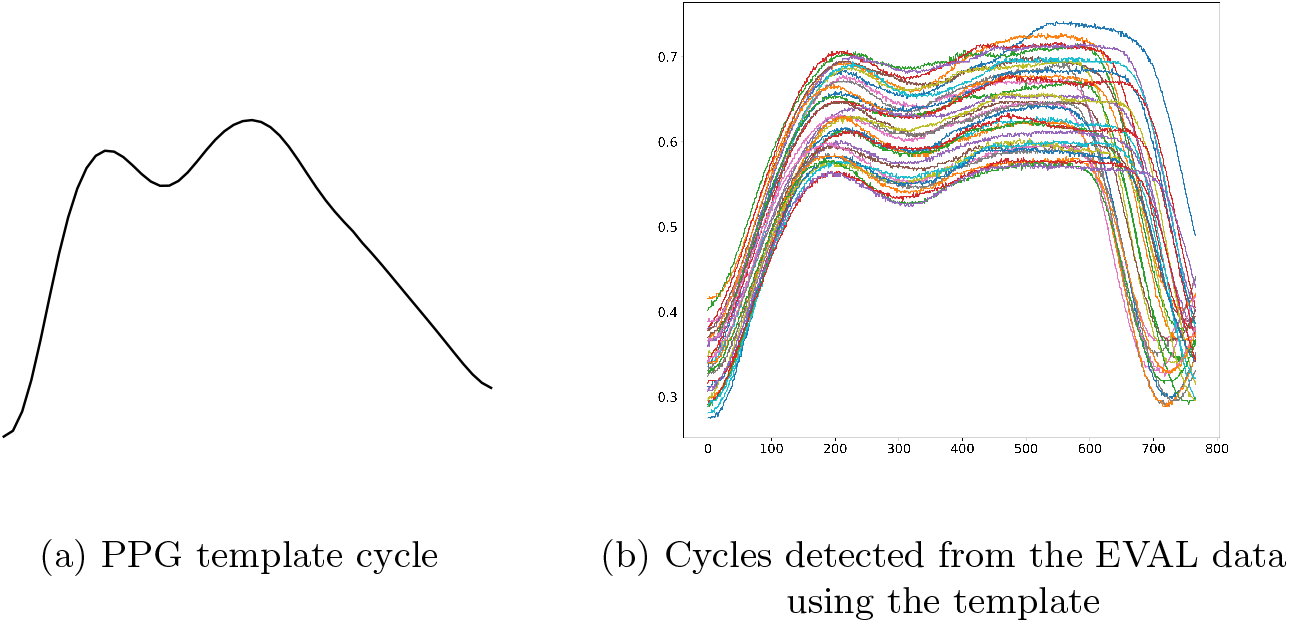
PPG cycles

## C Experiments

Here we briefly discuss how the different parameters of our experiments such as window size, motion filter etc. effect the results. In Figure 4 we can see the effect of adding motion filter in the stres test and 24-hours datasets. Though no prominent difference in MAE was seen during the controlled stress test, for the 24 hours dataset, using a motion filter clearly achieved better MAEs. In Figure 5 we can see the effect of different window sizes (length of the PPG signal considered as an input) on the results. In the stress test data the 15 sec and 30 sec window sizes performed considerably better than the 45 sec window size. In the 24-hours data the 30 sec and 45 sec windows performed better. Overall we chose the 30 sec window size for all our experiments. The last variable that we experimented with was the position of the window itself. Since the BP measurement happens over a duration of time there are different ways a window can be positioned. We particularly tested two: (1) Bfill, where the window starts exactly from the BP measurement start and goes t sec (t being the window size) before and (2) Bffill, where the window goes t/2 sec before and t/2 sec after the start of the BP measurement. In Figure 6 we can see the results. Here there was no clear winner between the two approaches.

**Fig. 4:**
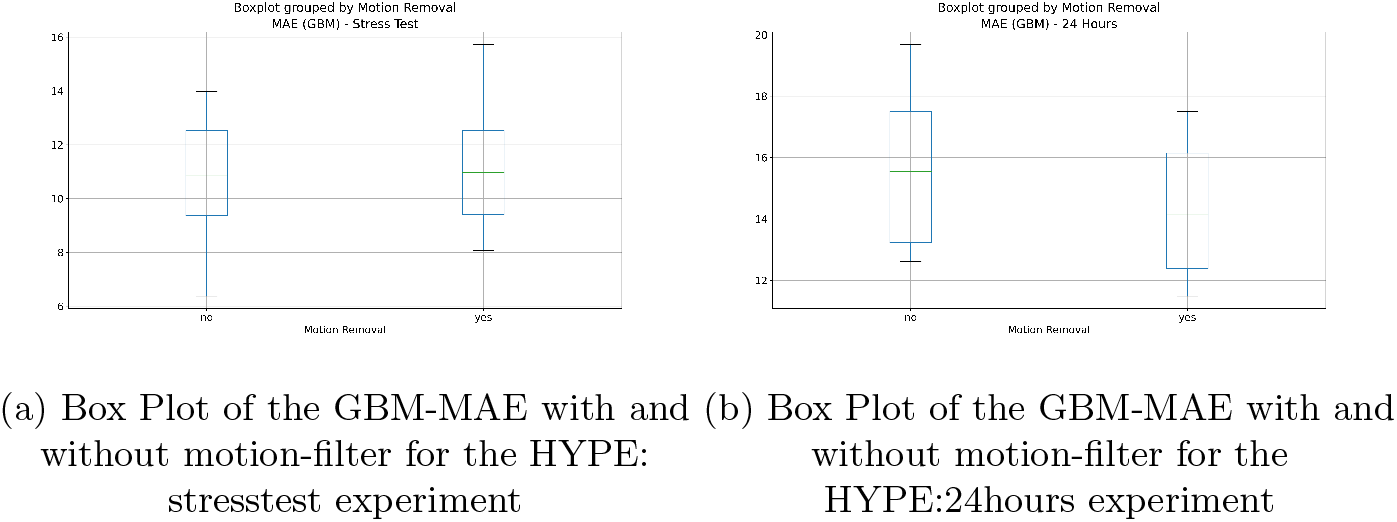
Box-plots depicting the effect of motion filters

**Fig. 5:**
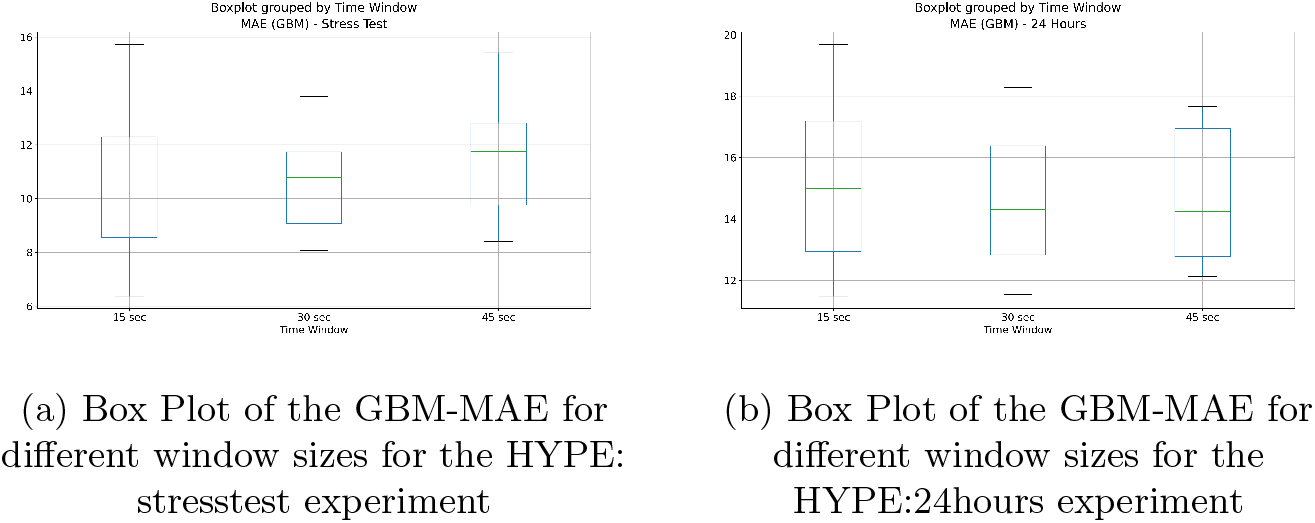
Box-plots depicting the effect of window sizes

**Fig. 6:**
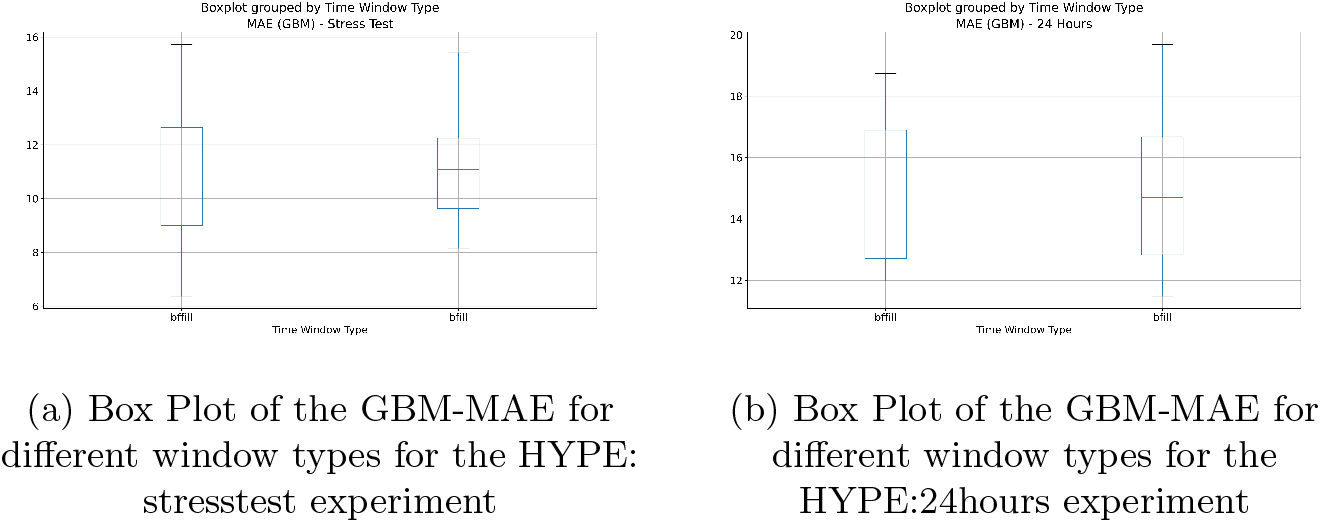
Box-plots depicting the effect of window types

